# High use of the emergency department shows typical features of complex systems: analysis of multicentre linked data

**DOI:** 10.1101/2020.10.08.20209296

**Authors:** Christopher Burton, Tony Stone, Phillip Oliver, Jon M Dickson, Jen Lewis, Suzanne Mason

## Abstract

**Objective:** High use of the ED is a worldwide problem. We hypothesised that high use of the ED could be understood as a feature of a complex system comprising patients, healthcare and society. Complex systems have characteristic statistical properties, with stable patterns at the level of the system emerging from unstable patterns at the level of individuals who make up the system.

**Methods:** Analysis of a linked dataset of routinely collected health records from all 13 hospital trusts providing ED care in the Yorkshire and Humber region of the UK (population 5.5 million). We analysed the distribution of attendances per person in each of three years and measured the transition of individual patients between high, low and non-attendance. We fitted data to power law distributions typically seen in complex systems using maximum likelihood estimation.

**Results:** The data included 3.6 million attendances at EDs in 13 hospital trusts. 29/39 (74.3%) analyses showed a statistical fit to a power law; 2 (5.1%) fitted an alternative distribution.. All trusts’ data fitted a power law in at least one year. Differences over time and between hospital trusts were small and partly explained by demographics. In contrast, individual patients’ high use was unstable between years.

**Conclusions:** ED attendance patterns are stable at the level of the system, but unstable at the level of individual high users. Attendances follow a power law distribution typical of complex systems. Interventions to address ED high use need to consider the whole system and not just the individual high users.

## Introduction

Meeting the needs of patients who are high users of urgent and emergency care is a major challenge for emergency medicine. High-using patients comprise a heterogeneous group with complex needs^1 2^, which for many comprise a mix of physical, mental and social problems^3 4^. While high use initially appears to a simple concept, it becomes less certain on closer inspection: definitions based on a threshold number of cases are abitrary^5^, definition of attendances for lower acuity problems^6 7^ is challenging, and the decisions patients make about use of emergency care are complex,^8^ reflecting personal and social factors^9 10^. Furthermore, interventions aimed at high-using patients, such as case management, which often appear effective at the level of the individual patient, appear less effective in controlled trials ^11 12^ leading some to argue that high use represents a downstream outcome of larger social systems.^13^

High use of the emergency department (ED) is a ubiquitous phenomenon^14^ but harbours a paradox: while high use is consistently found, the population of high-using patients is constantly changing. Studies in emergency medicine ^15 16^ have demonstrated that high use by an individual is a relatively unstable state: many high users in one time period become infrequent users in the next, and vice versa.

One way of understanding this paradox between the persistence, or stability, of high use in the ED and the transitory, or unstable, status of being a high user is to consider the problem from the perspective of a complex system^14 17^. Complex systems comprise many interacting components which “self-organise” rather than being externally directed or ordered^18^. Complex systems display order and stability at the level of the whole system which spontaneously emerges from apparent disorder and instability at the level of the system components. They also generate typical skewed distributions with a “heavy” tail containing high and very high values which typically follows a power law^19^. Supplementary data 1 describes power laws and the characteristics of power law distributions.

We hypothesised that a regional urgent and emergency care system would show typical features of a complex system: specifically, it would show a stable pattern of high use which followed a power law distribution.

## Methods

### Study Design

We carried out an analysis of routinely collected healthcare data for all ED care in the Yorkshire and Humber region of the UK, comprising 5.5 million residents over 3 consecutive years from April 2014 to March 2017.

### Patient and public involvement

No patients involved

### Data sources

We used a dataset extracted from the “Connected Health Cities: Data linkage of urgent care data” study (known as the “CUREd research database”) ^20^. This covers all EDs in the Yorkshire & Humber region. The CUREd research database contains over 23 million patient episodes of care between April 2011 and March 2017. Data relate to calls to the NHS111 telephone service; calls to the ambulance service; attendances at EDs, Urgent Care Centres and Minor Injury Units; and episodes of care within acute hospitals subsequent to an emergency admission. CUREd is a unique resource which enables the investigation of patient journeys across time, services and providers.

### Ethics and permissions

The CUREd database has approval from a National Health Service (NHS) Research and Ethics Committee, overseen by the NHS Health Research Authority’s Research Ethics Service, and from the NHS Health Research Authority (HRA), directly, to receive health and social care data without patient consent for patients of emergency and urgent care services in Yorkshire and Humber. The Leeds East REC granted approval (18/YH/0234) and, subsequent to receiving a recommendation to approve from the Confidentiality Advisory Group (18/CAG/0126, previously 17/CAG/0024), the NHS HRA provided approval for English health and care providers to supply identifiable patient data to the study. The study complies with the common law of duty of confidentiality owed by health professionals in regard to information provided by patients in the course of clinical care; the General Data Protection Regulation as enacted in the UK by the Data Protection Act 2018; and, where applicable, the Statistics and Registration Service Act 2007.

### Data extracted

The data used in the study comprised de-identified data extracted from routinely collected information on every ED attendance in the region, with all attendances for each individual patient linked by a single pseudonymised ID. We included all attendances at a type 1 ED (i.e a consultant led 24 hour service with full resuscitation facilities and designated accommodation for the reception of accident and emergency patients) by adults aged 18 or more at the time of attendance. Attendances were grouped by individual patient and by hospital trust. We used hospital trusts rather than individual EDs because they typically serve a distinct geographical population. While many trusts have only one ED, some trusts serve a conurbation with more than one ED and patients may visit either. Patients were categorised by age band (18-34, 35-54, 55-69, 70-84 and 85+), sex and by deprivation. The measure of deprivation was the Index of Multiple Deprivation (2015) for England. Patients were grouped into quintiles of deprivation relative to the entire English population. The distribution of patients by IMD quintiles in the data was not even; this reflects both the demography of Yorkshire and Humber generally – with more people living in deprived areas than the English average - and greater ED use by people of lower sociodemographic status.

### Analysis of characteristics and stability of ED use at system level

We carried out analyses at the level of the whole region (in order to examine the effects of age and socioeconomic status) and at the level of individual hospital trusts (to look for geographic variation). For each year we aggregated all attendances per patient and calculated the complementary cumulative distribution function, defined as the proportion of patients whose total number of attendances was equal to or greater than each number of attendances between 1 and the largest recorded. We then plotted this distribution with logarithmic axes. Plots showed data broken down by year and by either age band, socioeconomic status, or hospital trust. The technique of using logarithmic axes in this way means that a power law distribution appears as a straight line with a slope of one minus the scaling parameter. A larger power law scaling parameter indicates a steeper slope and a shorter tail to the distribution, while a smaller scaling parameter indicates a gentler slope and a longer tail.

We fitted power law distributions to data using maximum likelihood estimation with the poweRlaw package^21^ for R. We carried this out in four steps: inspection of plots; identification of best-performing minimum attendance number; fitting of distributions and estimation of confidence intervals. In step 1, we inspected plots of the data to find a plausible range of possible values for the minimum attendance number to use in the power law fitting (i.e. the number of attendances above which the shape of the distribution on logarithmic plots became linear). In step 2, we found the best-performing minimum attendance number by comparing the maximum likelihood fitting of the data to a power law starting at each value in the range of minimum attendance numbers from step 1. We then used this minimum attendance number as the lowest eligible number of attendances per patient for inclusion in the next two steps. In step 3, we tested the fit of the data to a power law using the Kolmogorov-Smirnoff test. We extracted the scaling parameter for the distribution and estimated p-values for the Kolmogorov-Smirnoff test by bootstrapping following the approach recommended in Clauset^22^ with 500 iterations. Where the p-value of the Kolmogorov-Smirnoff test was >0.05 we labelled the distribution as indistinguishable from a power law. When a distribution of data looked like a power law on the logarithmic plot but the p-value of the fit was <0.05 (i.e. the distribution differed significantly from a power law at some point) we compared the fit between a power law and two other distributions – the Poisson and lognormal - to find the distribution which best fitted the data. This comparison used a log likelihood ratio test ^22^. We labelled these distributions as similar to a power law. Finally, in step 4, we calculated 95% confidence intervals for each power law scaling parameter by bootstrapping with 400 iterations.

For the main analyses we used 12 month periods (April-March for each year in the data). We estimated power law scaling parameters for data split by year and additionally split by hospital trust, patient age or socioeconomic deprivation. We initially included all patients with complete data in each analysis but subsequently excluded patients over 70 from some of the analysis because the data for this group did not resemble a power law over the majority of the distribution. Finally we examined 6 month periods (beginning in April and October) in order to test for stability in the face of seasonal variation in demand.

## Results

Over the three years there were a total of 3,864,081 type 1 ED attendances. The total volume increased over the three years from 1,263,149 attendances (830,046 patients) in Year 1 (2014-15) to 1,310,167 (850,443 patients) in Year 3 (2016-17). This represents an increase in attendances and patients attending of 3.7% and 2.5% respectively between the first and third years.

The 13 hospital trusts varied substantially in size and demographics. Table 1 lists characteristics of each hospital trust including size of population served; number of ED patients; their median age and the percentage of patients in the most deprived quintile of the UK population. Some trusts covered a mix of urban and rural settings, while others served major conurbations with high levels of deprivation.

**Table 1.** Characteristics of hospital trusts and ED attenders

| Trust population served |  | ED patients in 2015-16 |  |  |
| --- | --- | --- | --- | --- |
| Hospital Trust | Number<sup>1</sup> (thousands) | Number (thousands) | Most deprived<sup>2</sup> | Median Age |
| A | 161 | 27 | 3% | 52 |
| B | 476 | 79 | 14% | 50 |
| C | 160 | 31 | 23% | 50 |
| D | 428 | 65 | 56% | 42 |
| E | 789 | 111 | 40% | 42 |
| F | 578 | 65 | 40% | 47 |
| G | 538 | 109 | 34% | 45 |
| H | 456 | 77 | 30% | 45 |
| I | 245 | 43 | 38% | 47 |
| J | 427 | 74 | 34% | 48 |
| K | 332 | 67 | 31% | 49 |
| L | 265 | 42 | 39% | 46 |
| M | 583 | 78 | 43% | 46 |

ED Emergency Department
<sup>1</sup> Estimated from NHS data for populations of corresponding clinical commissioning groups. This does not include patients who are seen at an ED but whose home address is outside the corresponding area.
<sup>2</sup> Proportion of ED patients whose postal code was in the most deprived 20% of the English population based on the Index of Multiple Deprivation 2015

### Visualisation of ED use at system level

Figure 1 shows the distribution of attendances per patient on logarithmic axes. Plots shown are for Year 2, but plots for each year are shown together in the upper part of supplementary figure 1. Figure 1a shows data points aggregated across all hospital trusts. The linear relationship between the log number of attendances and the log probability of a patient having that number or more – particularly between 3 and 30 attendances is indicative of a power law distribution. Figure 1b shows the data split by deprivation quintile. The gradient becomes less steep as deprivation increases. Figure 1c shows the data split by patient age: in this figure, the lines representing patients aged 70-84 and 85+ can be seen to curve differently from the straight lines of the other age groups. Figure 1d is a simplified version of Figure 1c with patients split into those aged under 70 years and those aged 70 and over. This highlights that the distribution for patients aged over 70 is convex on the logarithmic axes and does not have the linear appearance of a power law until a minimum attendance number of around 10. This distribution has a shorter tail. In summary, for all groups except patients aged over 70 individual attendance patterns appeared on visual inspection to fit a power law.

**Figure 1.**
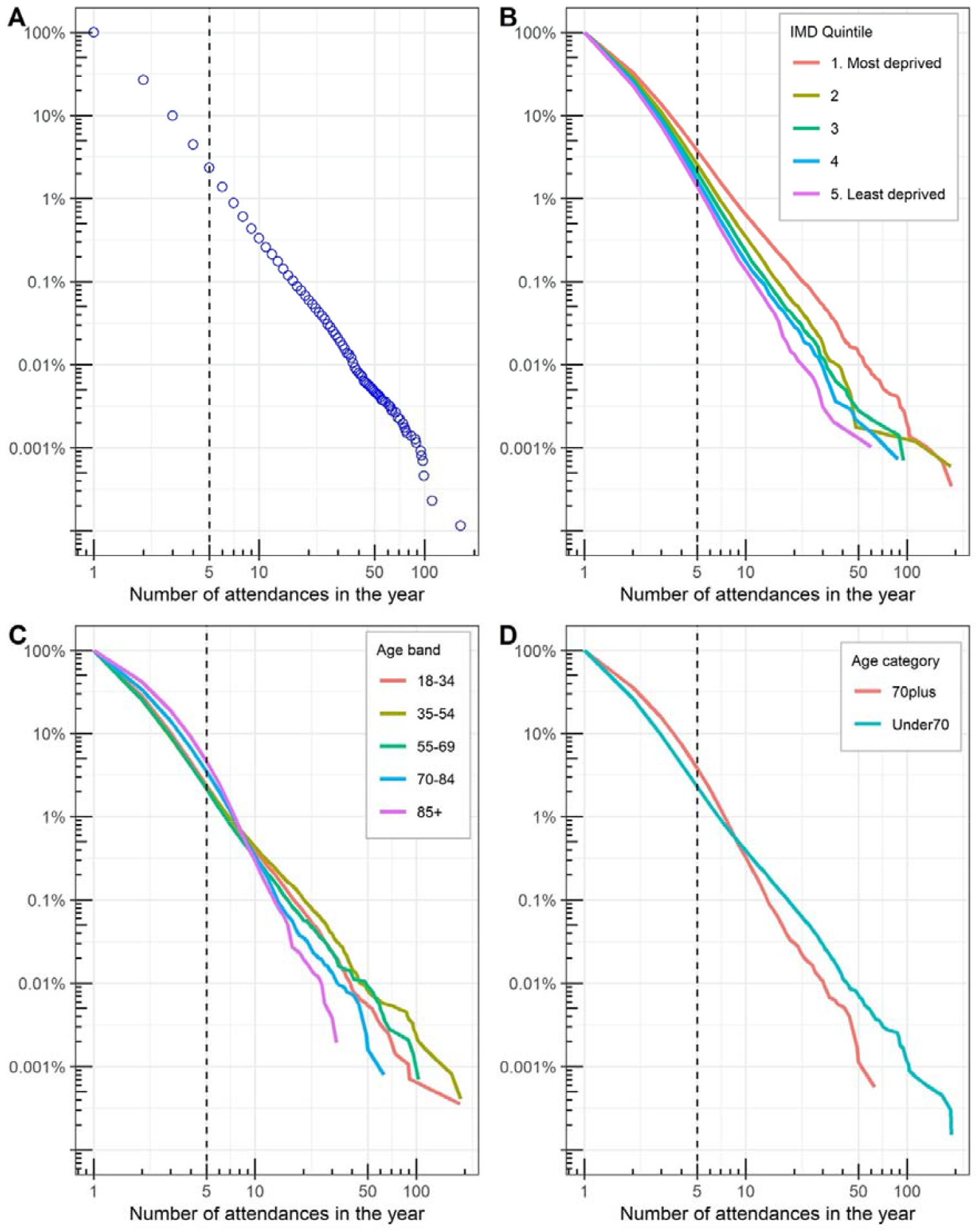
Distributions of ED attendance. Axes represent number of attendances in a year and the proportion of patients ED attenders who make at least that number of attendances. Figure 1A represents all patients in one year (using points to indicate spacing of values), Figure 2B shows data split by socioeconomic deprivation status, and Figure 2C by age group. Figure 2D is a simplified version of 2C. Figures 2B-2D use lines rather than points to reduce crowding of the data.

### Estimation of power law fitting

From observation of the data and preliminary testing of model fit, we found that the best-performing minimum value of attendance for power law fitting was 3; this value was used in all subsequent model fitting. Table 2 shows the results of analysis of power law fitting at the level of year and hospital trust. The data were indistinguishable from a power law in 29/39 instances. Of the remaining 10 instances, 8 were similar to a power law (they were a better fit to a power law than a Poisson distribution and there was no difference in fit between a power law and a lognormal distribution). The remaining two instances showed better fit to a lognormal distribution. They were from the same trust, but data from that trust in the remaining year was indistinguishable from a power law. Plots of data from this trust (M) show minor deviations from a power law in years 1 and 3 but the overall pattern is still clearly heavy-tailed (lower part of supplementary figure 1). Data pooled across trusts was indistinguishable from a power law in 7/15 analyses split by year and deprivation quintile and 3/9 analyses split by year and age group. In all remaining analyses, data were similar to a power law. As 61/63 distributions analysed were indistinguishable from, or similar, to a power law distribution we included all of them in comparisons of power law scaling parameter.

**Table 2.** Fit of power law distribution to data from each hospital trust by year

| Hospital Trust | Year 1 |  |  |  | Year 2 |  |  |  | Year 3 |  |  |  |
| --- | --- | --- | --- | --- | --- | --- | --- | --- | --- | --- | --- | --- |
|  | Scaling Parameter<sup>1</sup> | KS<sup>2</sup> p-value | LRT<sup>3</sup> p-value | Distribution | Scaling Parameter | KS p-value | LRT p-value | Distribution | Scaling Parameter | KS p-value | LRT p-value | Distribution |
| A | 3.65 | 0.09 |  | Power Law | 3.65 | 0.07 |  | Power law | 3.59 | 0.14 |  | Power law |
| B | 3.56 | 0.23 |  | Power Law | 3.52 | 0.00 | 0.53 | Uncertain | 3.46 | 0.08 |  | Power law |
| C | 3.66 | 0.46 |  | Power Law | 3.84 | 0.30 |  | Power law | 3.73 | 0.82 |  | Power law |
| D | 3.41 | 0.23 |  | Power Law | 3.41 | 0.12 |  | Power law | 3.40 | 0.10 |  | Power law |
| E | 3.37 | <0.01 | 0.38 | Uncertain | 3.35 | 0.02 | 0.50 | Uncertain | 3.34 | 0.19 |  | Power law |
| F | 3.48 | 0.02 | 0.10 | Uncertain | 3.41 | 0.66 |  | Power law | 3.29 | 0.23 |  | Power law |
| G | 3.65 | 0.01 | 0.89 | Uncertain | 3.63 | 0.00 | 0.35 | Uncertain | 3.63 | 0.04 | 0.17 | Uncertain |
| H | 3.67 | 0.38 |  | Power law | 3.59 | 0.69 |  | Power law | 3.52 | 0.38 |  | Power law |
| I | 3.57 | 0.26 |  | Power Law | 3.59 | 0.33 |  | Power law | 3.56 | 0.18 |  | Power law |
| J | 3.66 | 0.29 |  | Power law | 3.59 | 0.63 |  | Power law | 3.62 | 0.29 |  | Power law |
| K | 3.46 | 0.10 |  | Power Law | 3.48 | 0.47 |  | Power law | 3.49 | 0.86 |  | Power law |
| L | 3.55 | 0.04 | 0.26 | Uncertain | 3.49 | 0.14 |  | Power law | 3.59 | 0.85 |  | Power law |
| M | 3.48 | <0.01 | <0.01 | Lognormal | 3.46 | 0.08 |  | Power law | 3.48 | 0.03 | 0.02 | Lognormal |

<sup>1</sup> Scaling parameter for power law fit for all patients with 3 or more attendances.
<sup>2</sup> Kolmogorov Smirnov test p-value (if >0.05 indicates data indistinguishable from a power law)
<sup>3</sup> Likelihood Ratio Test p-value (only applied if data not indistinguishable from a power law; if >0.05 indicates no difference in fit between lognormal and power law distribution)

Table 3 summarises three measures of ED attendance by deprivation quintile and age group. The total numbers of attendance for deprivation reflect both increased prevalence of socioeconomic deprivation in the region (quintiles are for the whole population of England not just Yorkshire and Humber) and increased ED use by the most socioeconomically deprived. The power law scaling parameter is inversely related to socioeconomic deprivation. In terms of the plots in figure 1 a smaller scaling parameter equates to a shallower slope, meaning that the probability of a patient having a given number of attendances is higher, and the probability of having no further attendances is lower. The confidence intervals around the power law scaling parameters in table 2 indicate that the differences between deprivation levels within the same year are unlikely to have arisen by chance.

**Table 3.**
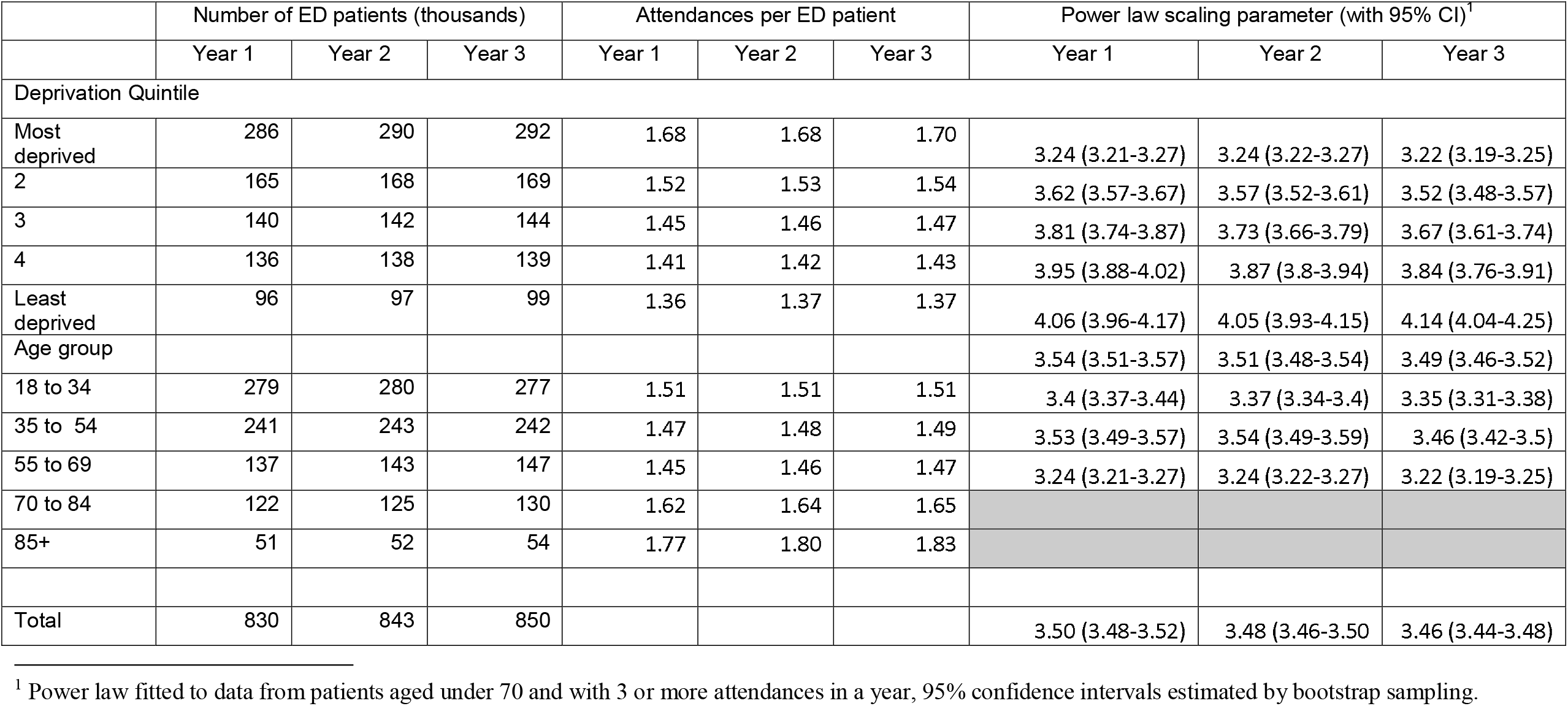
ED attendance characteristics by population deprivation quintile and age group

Table 3 also permits assessment of trends in ED attendance. The number of attendances increased over time in all subgroups apart from those aged 18-34. The power law scaling parameter decreased (i.e., greater probability of high attendance) except for the highest and lowest socioeconomic deprivation groups. The finding of little change in the most deprived is surprising given that much of the perception about ED capacity has focused on unnecessary attendance in this group.

Figure 2 examines the variation in power law scaling parameters between hospital trusts. Figure 2a shows the variation between years within trusts. From this it appears that three trusts (F,B & H) show more marked year to year change with the scaling parameter reducing over time (i.e. towards greater reattendance). Two of these three trusts served less deprived populations, but as the scaling parameters exclude patients over 70 this could not simply be due to increasing numbers of older patients. In figure 2b the scaling parameter for a single year (Year 2) is plotted against the proportion of patients in the most deprived population quintile. It suggests that at least part of the variation between trusts is attributable to population differences. Analysis in shorter periods found no consistent seasonal pattern between semesters (supplementary figure 2).

**Figure 2.**
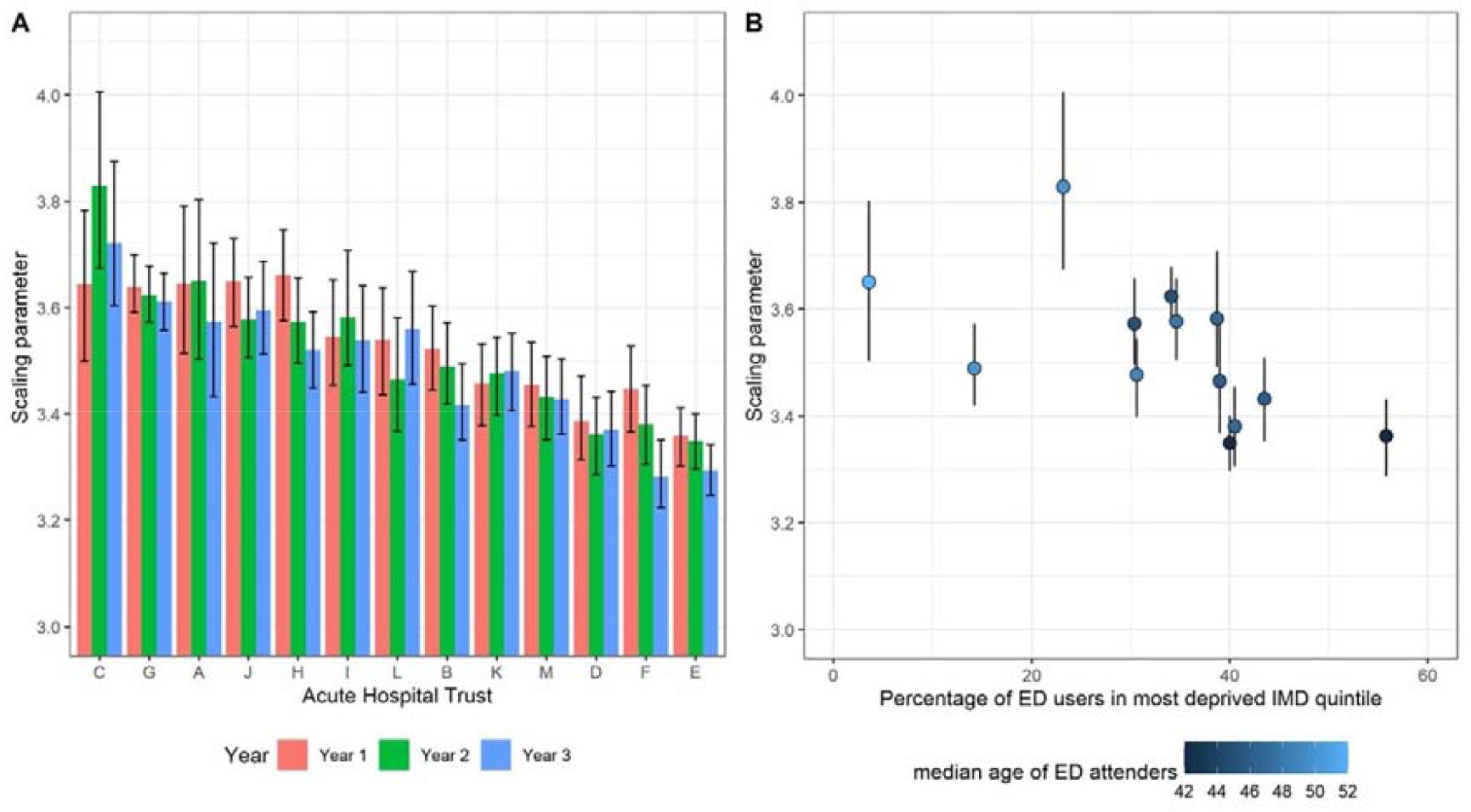
Variation in power law scaling parameter by hospital trust. Figure 2A shows the year to year variation for each hospital trusts ordered by scaling parameter. Figure 2B shows the relationship between power law scaling parameter, socioeconomic deprivation and median patient age for each hospital trust. Error bars in both plots indicate 95%confidence intervals.

## Discussion

### Summary of principal findings

This study confirmed the hypothesis that ED attendance patterns follow power law distributions. These findings were consistently present across 13 hospital trusts serving a population of 5.5 million and suggest that Emergency Department use behaves as a complex system.

### Limitations

Despite the size of population served it is possible that some of the features we observed were local rather than general phenomena, however the consistency of findings across a very socioeconomically diverse region suggests a generalisable process. While data did not provide a precise fit to a power law in every analysis – particularly when aggregating across hospital trusts, the absence of a better fitting distribution in almost all cases suggested that this lack of fit may be explained by local ‘noise’ in the data rather than a fundamental misapplication of the model.

### Relationship to other research

A review of published studies to 2017 documented heavy tailed distributions in use from over 20 EDs but only one tried fitting a power law distribution^14^. None of these studies was large enough to examine the impact of demographic features on these patterns. The study places research into ED high use alongside a wider body of quantitative work about complex systems. While complex systems science is increasingly contributing to other areas of medicine^23^, it has only rarely been used to address pressing problems of health system use^24^.

### Implications

The approach we have used for fitting power laws and related distributions to ED data has implications for measurement and for understanding of the problem of high use. The fitted parameters provide an objective measure by which to quantify the dynamics of ED use. This can be used in conjunction with statistical control methods to provide a means for identifying when and under what circumstances systems may change, or fail to follow this distribution, including in evaluating new interventions to manage demand.

Thinking of ED use as a complex system has important implications: first any evaluation of interventions to reduce high use in the ED needs to take a whole system view^25^. While a solution for an individual may benefit that person, if it simply means that another patient occupies their place in the power law distribution of attendance, then the emergency medicine system will be no better off. Second, the very stability of complex systems, which we have demonstrated in ED use, makes them challenging to change. Numerous initiatives have sought to reduce demand on the ED. As interventions in complex systems, one would expect most interventions addressing ED high use to have small effects – to be buffered by the system – but one would also expect a few interventions to have larger effects ^26^, potentially leading to pressure to adopt them elsewhere with limited success. Third because ‘high’ use can be seen to be just one part of a continuous spectrum of use, strategies to reduce reattendance should consider the effects of processes which occur in many consultations: these may include defensive safety netting (“come back if you have any concerns”) and unthinking emphasis on patient satisfaction (much of which derives from business models designed to generate ongoing demand). Finally, the concepts and analytic tools used here, which are widely established in other fields, can be readily transferred to the evaluation of healthcare.

## Conclusion

This study found compelling evidence that high use of the emergency department can be understood as a complex system. The concepts and analytic tools used here can be used to design, evaluate and model interventions to address high use, in order to ensure that they do more than replacing one high-using individual with another.

## Supporting information

suppplementary data

STROBE

## Data Availability

Data are not available for sharing.

