## Supplementary material for "High use of the emergency department shows typical features of complex systems: analysis of multicentre linked data": suppplementary data

Supplementary data

**Power law distributions**

In a power law distribution, the probability that an event of magnitude *X* will occur follows the equation P_(_*_X_*_)_=*kX ^-α^* where *k* is a constant and *α* is termed the power law scaling parameter. In addition, the probability P_(x≥_*_X_*_)_ that an event of magnitude of at least *X* will occur approximates to *kX^(1-α)^.* This empirical cumulative distribution function can readily be plotted and if logarithmic axes are used, produces a straight line. In real-world data power laws typically only occur at values of X above a threshold.

In the context of high use of the ED, P_(_*_X_*_)_ refers to the probability of a patient having *X* attendances and P_(x≥_*_X_*_)_ the probability of a patient having at least *X* attendances.

Power law distributions typically have a median value of 1 and a mean value of close to 1. Probabilities diminish quickly as the magnitude of *X* increases (for instance with k=1, *α*=3 and *X* = 4 then P_(_*_X)_* = 0.015 and P_(x≥_*_X_*_)_ = 0.0625 and for *X*=10, P_(_*_X)_* = 0.001 and P_(x≥_*_X_*_)_ = 0.01). Despite this, power law distributions of sufficient size still include very large values for X: for instance using the scaling parameter above the probability of a value for *X* of at least 100 is 100^-2^ or 1:10,000. This occurrence of values within the distribution which are far from the median accounts for the observed “heavy tail” of the distribution.

Power laws can be fitted to empirical data using maximum likelihood estimators and their scaling parameters estimated. Fitting power laws to different sets of data means their scaling parameters which describe the distribution can be compared objectively.

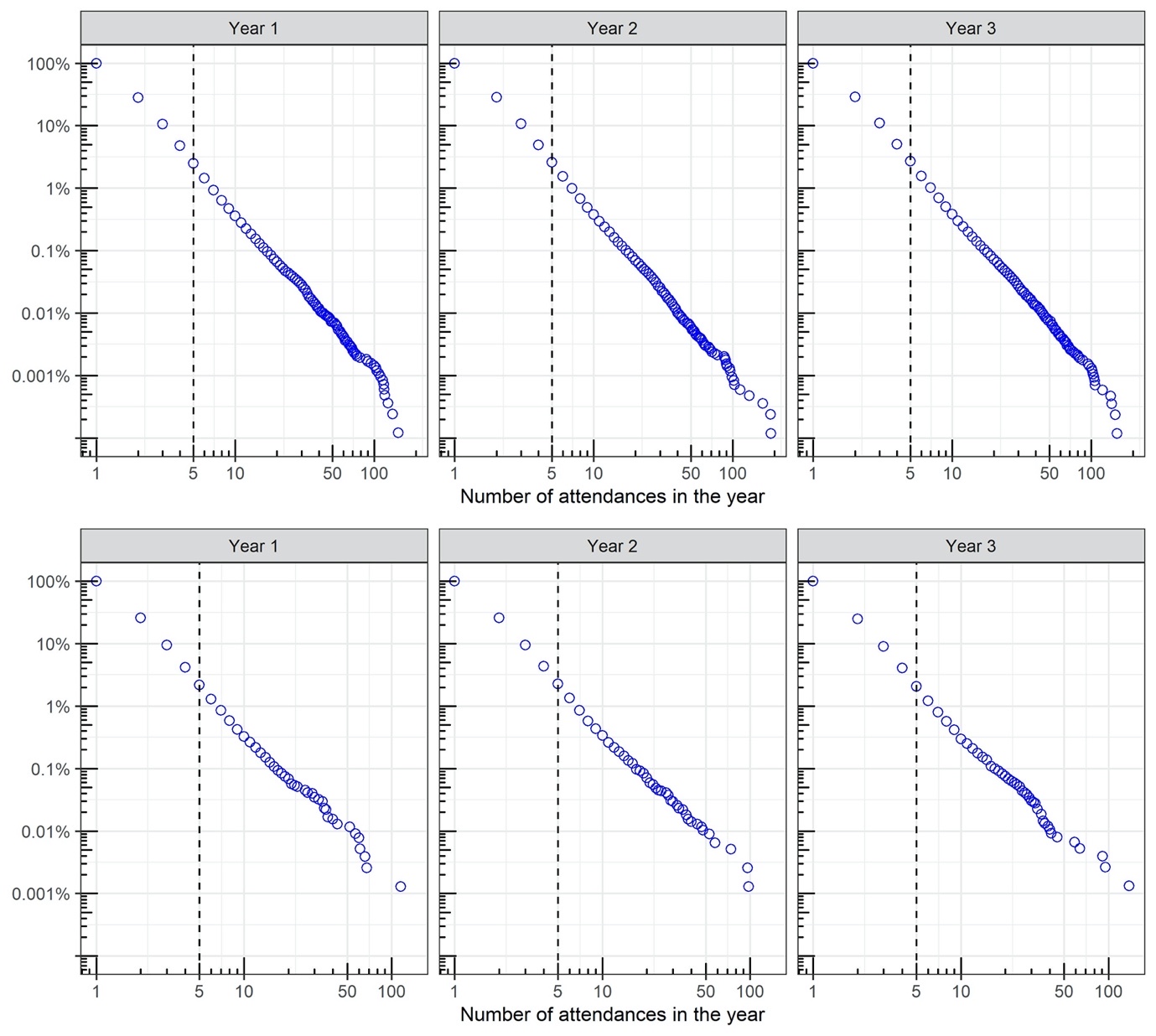

Supplementary Figure 1 **Distributions of ED attendance by year.** Upper row is for data for all patients across Yorkshire & Humber, lower row is for data from practice M indicating the two years (Year 1 and Year 3) which did not fit a power law. Axes represent number of attendances in a year and the proportion of patients ED attenders who make at least that number of attendances.

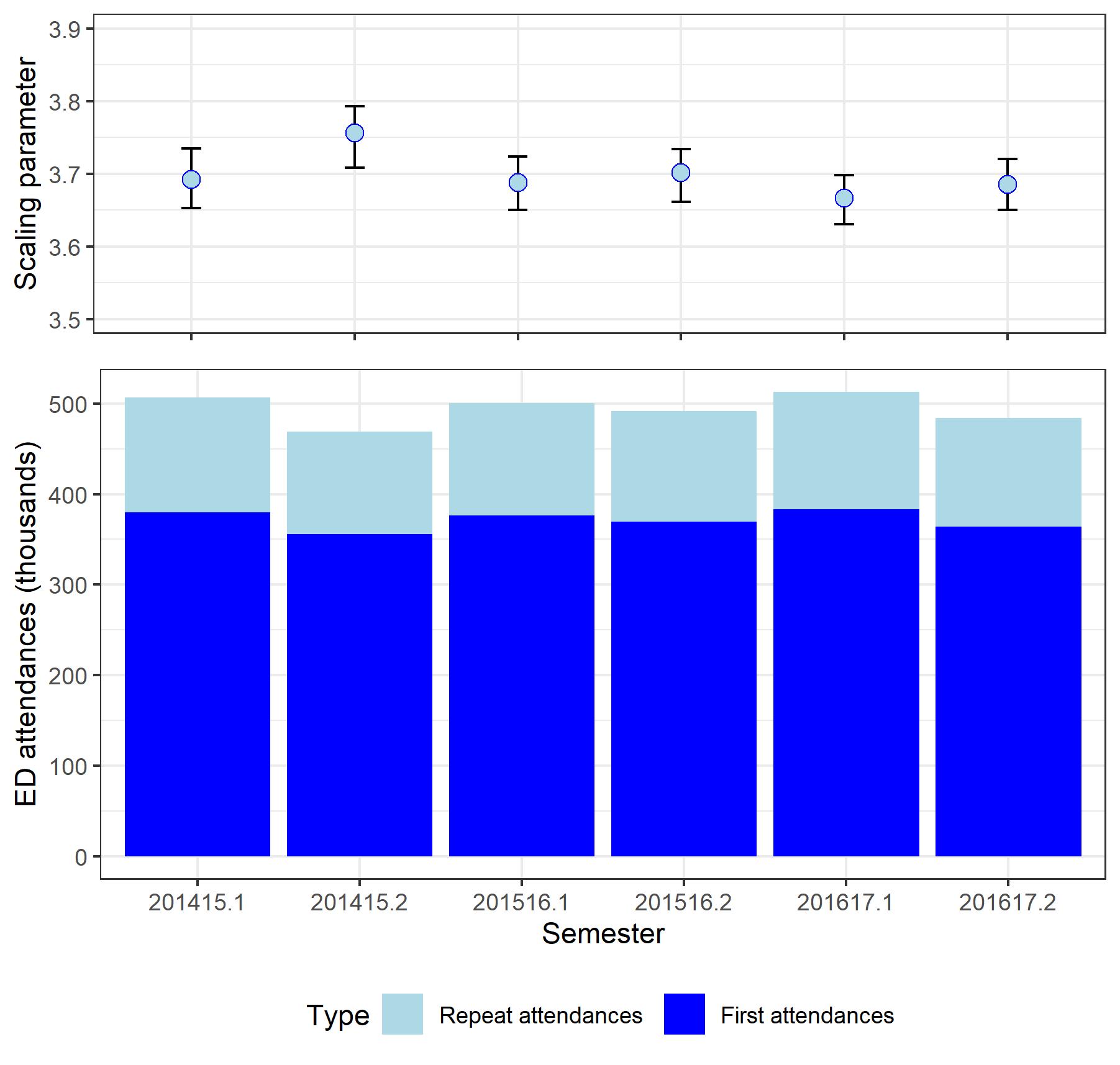

**Supplementary Figure 2 Changes in power law scaling parameter and attendances by 6 month period**
Semester 1 is April to September and semester 2 October to March. Lower plot shows number of ED attendances (split into first and repeat) in each period and upper plot shows the corresponding power law scaling parameter.
